# Methodological updates in the Conventional Gait Model 2 preserve kinematic reliability in asymptomatic and cerebral palsy individuals

**DOI:** 10.64898/2026.01.12.26343932

**Authors:** F. Leboeuf, M. Sangeux, M Fonseca, C. Dussault-Picard, S. Armand

## Abstract

Three-dimensional gait analysis is widely used to support clinical decision-making in neuromuscular disorders, with the Conventional Gait Model (CGM) being the most commonly applied biomechanical model in clinical practice. Recent developments of the CGM, grouped under the open-source CGM2 framework, introduced methodological updates intended to improve robustness while preserving backward compatibility. However, the reliability of these successive CGM2 iterations has not been comprehensively evaluated, particularly in pathological gait populations.

This study investigated within- and between-assessor reliability of lower-limb kinematics across three CGM2 versions (2.1, 2.2, and 2.3) in asymptomatic participants and individuals with cerebral palsy. Reliability was quantified using standard error of measurement and minimal detectable change across the gait cycle. Overall measurement error remained low and consistent across models and participant groups, with standard errors close to 2° and minimal detectable changes around 6°. Introducing kinematic fitting had minimal influence on reliability, while adding tracking markers on the thigh and shank produced a modest reduction in hip transverse rotation error.

These findings indicate that methodological refinements implemented in CGM2 preserve the reliability of the original CGM while providing incremental improvements for clinically relevant parameters, supporting its use in both asymptomatic and pathological gait analysis and longitudinal clinical assessments

## 1. Introduction

Three-dimensional gait analysis plays a central role in the clinical management of neuromuscular disorders affecting human locomotion [1,2]. Since its introduction in the early 1990s, the Conventional Gait Model (CGM) [3–5] has become the most used model for computing joint kinematics and kinetics from marker-based motion capture data in clinical practice [6]. Combined with the extensive clinical experience accumulated over the years, reliability studies of the CGM [7] have strengthened clinicians’ understanding of the model’s approximations and increased confidence in using 3D gait analysis to support diagnosis, guide therapeutic interventions, and monitor patient outcomes [8].

Despite its widespread use, the CGM has not been adopted as a community standard for clinical gait analysis. On one hand, alternative models, such as the IOR [9] or CAST [10], HBM [11] are also used in some laboratories [6] Furthermore, some gait laboratories have developed their own implementations to more easily integrate advances in biomechanical modelling [6]. On the other hand, the CGM relied on inaccurate methods, particularly in locating hip joint centres [12] or tracking bone motion [13]. Using fluoroscopy as a reference, Leboeuf et al. [13] demonstrated that the original segmental pose method [3]—based solely on marker trajectories without mathematical optimization—did not reproduce transverse plane rotations with an accuracy better than 5°.

Recognizing the limitations of the original CGM, an open-source initiative [14], named CGM2 (www.pycgm2.netlify.app) was developed to reproduce the original CGM outputs while introducing stepwise methodological improvements. Independent studies by Leboeuf et al. [14] then Kerr et al. [15] confirmed that CGM2 faithfully replicated the kinematics of the original model in typically developing children. CGM2.1 modified the computation of the hip joint centre by implementing Hara’s regression equation [16]. Subsequent iterations, CGM2.2 and CGM2.3, introduced more substantial technical changes, including the use of kinematic fitting (a.k.a inverse kinematics) as the default method for defining segmental pose, and the replacement of traditional wand-mounted markers with additional skin clusters on the thigh and shank [17]. This transition from wand-mounted markers to skin-mounted clusters is expected to enhance the repeatability of kinematic outputs, as Fonseca et al. [18] demonstrated through a sensitivity analysis that wand markers are a major source of error.

To date, few studies have investigated the repeatability of the CGM2 in gait and none focused on pathological gait populations such as individuals with cerebral palsy (CP). Outside its primary application to gait, McAllister et al. [19] and Okahisa et al. [20] reported good to excellent between-day reliability (Intra correlation class > 0.75) for lower-limb kinematics and kinetics during running and cycling, respectively. Horsak et al. [21] conducted the only study focusing on gait, specifically in participants with obesity. Using the Standard Error of Measurement (SEM)—as recommended by McGinley et al. [7] and the COSMIN consensus [22] for assessing measurement error— Horsak et al. [21] reported acceptable test–retest reliability during level walking and stair negotiation, with SEM ranging from 2 to 5°. While these findings support the use of CGM2 from a reliability perspective, additional work is required to facilitate its adoption in clinical gait analysis. Reliability should be established in pathological populations typically referred for gait assessment, such as participants with cerebral palsy.

The aim of this study was (1) to quantify the within- and between-assessor reliability with the methodological changes introduced in the CGM2 variants, and (2) to determine whether these changes improve the reliability of kinematics in participants with cerebral palsy (CP) and asymptomatic participants. Specifically, the transition from CGM2.1 to CGM2.2 was designed to assess the effect of the segmental pose method, whereas the transition from CGM2.2 to CGM2.3 aimed to evaluate the effect of the marker set modification.

## 2. Method

### 2.1. Participants

Thirty-eight participants aged between 6 and 43 years were recruited from the Geneva University Hospitals. Participants were divided into the cerebral palsy (CP) group, when a confirmed diagnosis of CP have been established (n=19) and the asymptomatic group, when they had no pathological conditions affecting normal motor abilities (n=19). According to the Gross Motor Function Classification System (GMFCS), the CP group was composed of GMFCS level I: n = 15, GMFCS level II: n = 3, and GMFCS level III: n = 1. Anthropometric data and characteristics for both the asymptomatic and CP groups are presented in Table 1. The research protocol was approved by the “Commission Cantonale d’Éthique de la Recherche Genève” (CCER-2020-00358), and all participants, or their legal guardians in the case of minors, provided written informed consent.

**Table 1.**
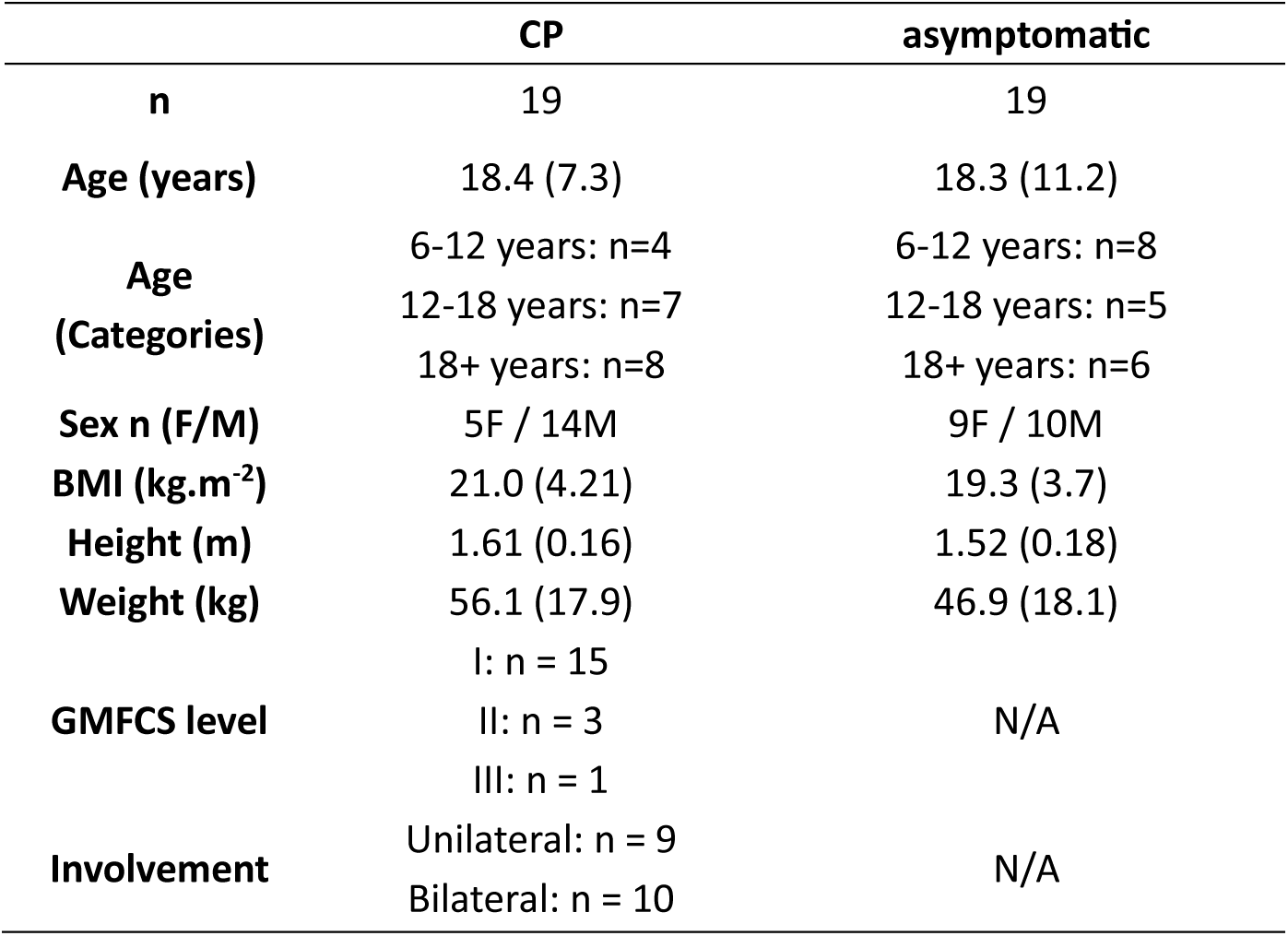
Mean (standard deviation) of participant anthropometrics and characteristics for the cerebral palsy (CP) and the asymptomatic groups. Sample size (n), side of the muscle disorder involvement; unilateral or bilateral, or not applicable (N/A), and sex; female (F) and male (M) are also reported.

|  | CP | asymptomatic |
| --- | --- | --- |
| n | 19 | 19 |
| Age (years) | 18.4 (7.3) | 18.3 (11.2) |
| Age (Categories) | 6-12 years: n=4<br>12-18 years: n=7<br>18+ years: n=8 | 6-12 years: n=8<br>12-18 years: n=5<br>18+ years: n=6 |
| Sex n (F/M) | 5F / 14M | 9F / 10M |
| BMI (kg.m<sup>-2</sup>) | 21.0 (4.21) | 19.3 (3.7) |
| Height (m) | 1.61 (0.16) | 1.52 (0.18) |
| Weight (kg) | 56.1 (17.9) | 46.9 (18.1) |
| GMFCS level | I: n = 15<br>II: n = 3<br>III: n = 1 | N/A |
| Involvement | Unilateral: n = 9<br>Bilateral: n = 10 | N/A |

### 2.2. Experimental protocol

All participants visited the gait laboratory on two occasions within a 10-day interval to minimize potential changes in gait due to disease progression or anthropometric modifications. Passive reflective markers (14 mm) were placed according to the CGM specifications (table 2), with palpation following the previously established guidelines [23]. During the first visit, each participant underwent a gait analysis session conducted only by assessor A, who was responsible for marker placement. In the second visit, each participant completed two new gait analysis sessions, each conducted by a different assessor (A and B). Both assessors had received prior training in marker placement and gait analysis procedures. To avoid bias in marker placement, only the assessor in charge of the respective session was present during marker positioning.

**Table 2.**
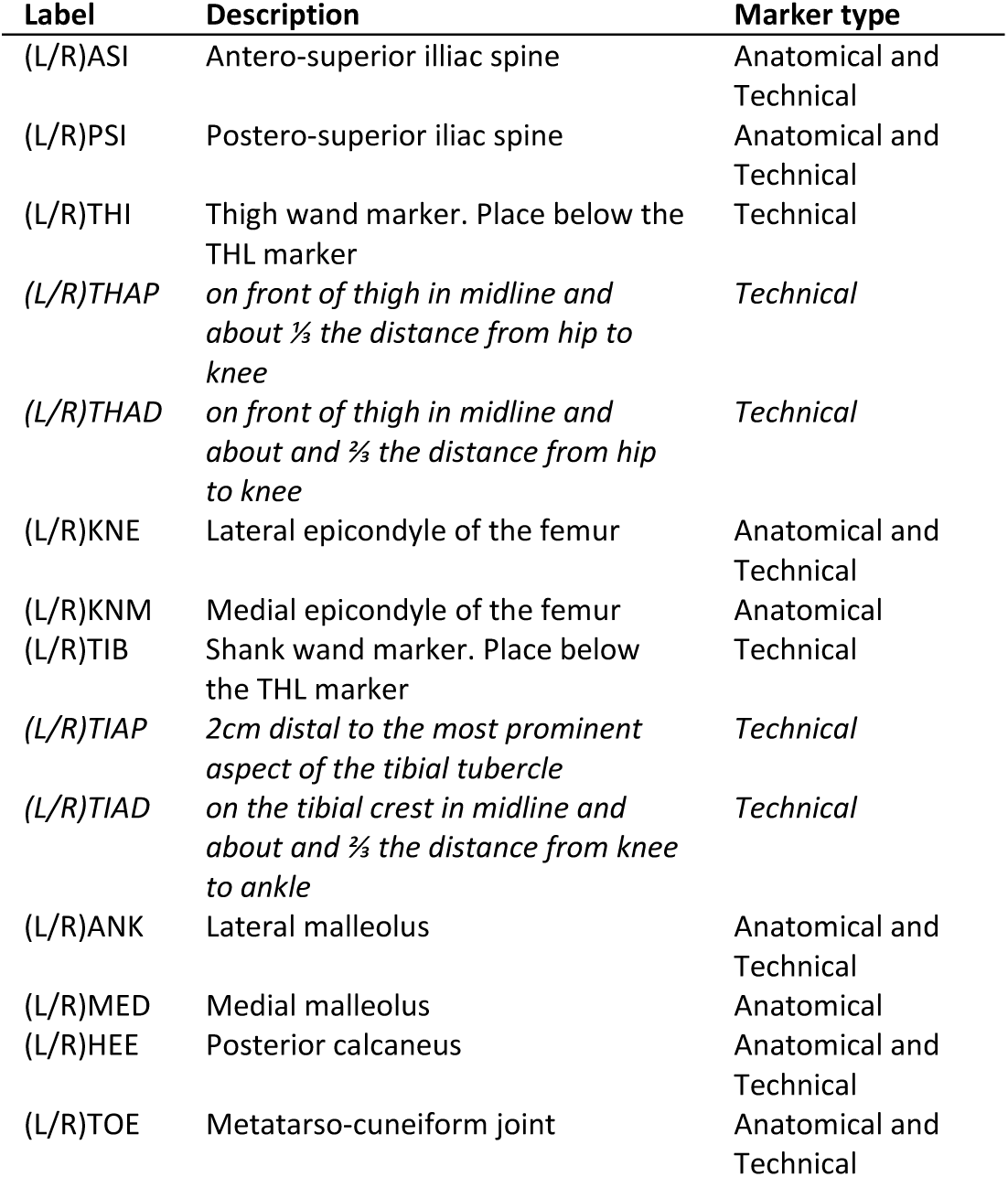
Description of the marker location. *Italic* markers are the additional markers of the CGM2.3 only

Participants were instructed to perform one static trial and multiple walking trials barefoot to obtain at least 10 gait cycles. Marker trajectories were tracked using a 12-camera motion capture system (Oqus7+, Qualisys, Göteborg, Sweden) at 100 Hz. Anthropometric data for each participant were measured by assessor B during both visits.

### 2.3. Gait modelling and processing

All analyses were conducted using the open-source library pyCGM2 (www.pycgm2.netlify.app), which allows seamless implementation of the different iterations of CGM2 (2.1, 2.2, and 2.3) on the same gait trial. CGM2.1 retains the original marker set of the CGM, as well as the original method for calculating joint orientations, which is based on tracking marker trajectories at each captured frame. In contrast, CGM2.2 performs pose estimation with kinematics fitting through OpenSim’s optimization algorithm [24]. CGM2.3 also employs kinematic fitting but uses additional tracking markers placed on the thigh and tibia.

Despite these differences, all three iterations of the CGM share the same definitions for joint centers and axes. The hip joint center is calculated using Hara’s regression equation [16], while the knee joint center is defined as the midpoint between the lateral and medial condyles, determined during the static trial. No correction for knee varus-valgus was applied in the models used, ensuring consistency in the comparison of results across the different iterations.

### 2.4. Data processing

The within-assessor standard error of measurement (SEM-WA) and between-assessor standard error of measurement (SEM-BA) were calculated using an ANOVA, where sessions and assessors were considered as explanatory variables. Both lower limbs were grouped together for asymptomatic participants, whereas only the affected lower limb was retained for participants with CP. The upper 95% confidence limit (CL) was calculated assuming the variances follow a chi-squared distribution, with the CL expressed as a multiplier of the SEM (as described by Stratford et al. [25]):

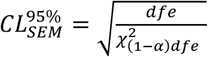

where *dfe* is the number of degrees of freedom. *dfe* = *n*_*p*_(*n*_*m*_ − 1) with *n*_*p*_ is the number of participants and *n*_*m*_ is the number of measurements per participant, and 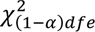 is the chi-squared distribution for a probability level of a and the same number of degrees of freedom. As there is no obvious clinical significance to the lower CL, a value of a = 0.05 was used rather than 0.025, as used by Stratford et al. [25].

The minimal detectable changes (MDC) were computed as:

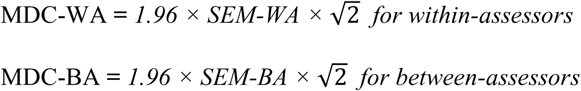

Additionally, the ratio (*r*) representing SEM-BA divided by SEM-WA was calculated as an index of assessor-related measurement error. Reliability was assessed using the lme4 package (v1.1-27.1) available in R (R Core Team 2020).

## 3. Results

### 3.1. Global and segmental SEM across gait models

The overall SEM (table 3) ranged from 1.4° to 2.3°, regardless of the group and CGM iterations. Similarly, overall MDC values fluctuated between 3.9° and 6.3°. Both SEM-BA and MDC-BA were slightly higher compared to SEM-WA and MDC-WA. The largest increase in MDC was observed in the asymptomatic group using CGM2.3, where the MDC-WA was of 3.9° and MDC-BA of 5.6°.

**Table 3.**
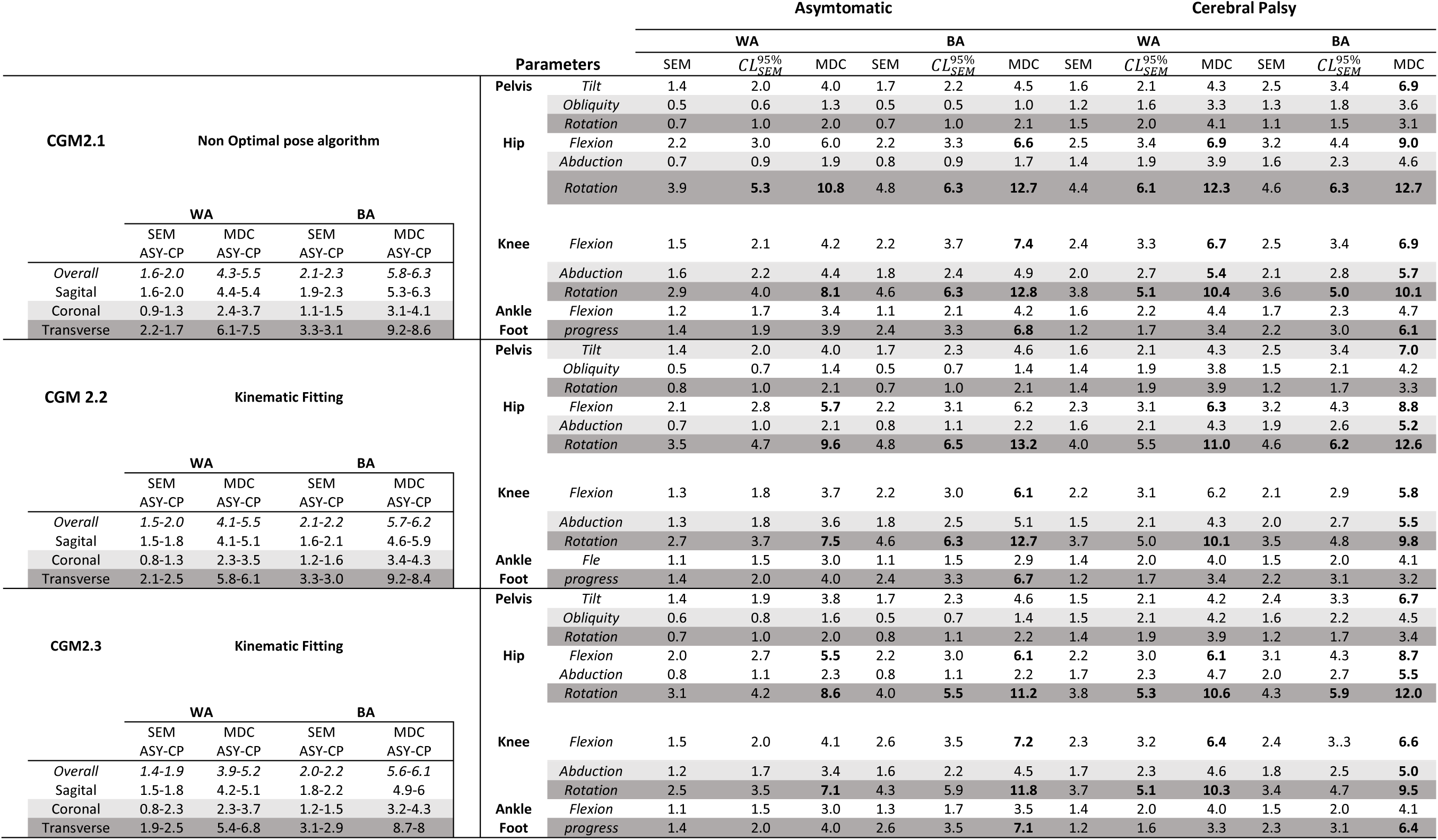
Average between-assessors (BA), within-assessor (WA) standard error of measurement (SEM), and minimal detectable change (MDC) calculated for the asymptomatic (ASY) and cerebral palsy (CP) groups, and across conventional gait models CGM2.1, CGM2.2 and CGM2.3 The upper 95% confidence limit 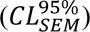 was calculated under the assumption that the variances follow a chi-squared distribution. Bold indicates value over the 5° threshold.

Concerning SEM and MDC across different segmental planes (figures 2 and 4), the highest values were observed in the transverse plane. The maximum SEM-BA and MDC-BA were recorded in both asymptomatic and CP participants using CGM2.1, with MDC-BA values reaching 9.2° and 8.6°, and SEM-BA reaching 3.3° and 3.1°, respectively. Similarly, CGM2.1 also exhibited the highest SEM-WA and MDC-WA, with 7.5° and 1.7° for CP and 6.1° and 2.2° for asymptomatic participants.

Figures 1 and 3 illustrate the frame-by-frame profiles of the SEM and the ratio. Overall, ratios were greater than 1 across all CGM2, indicating higher SEM-BA than SEM-WA. Regardless of the CGM2 iteration, both the CP and asymptomatic groups showed (i) the highest ratio (*r*>2) for foot progression angle and (ii) an increase in hip rotation and knee abduction SEM-WA during early swing, occasionally exceeding thse 5° threshold. In the asymptomatic group (figure 1), knee flexion SEM-BA values were noticeably higher than SEM-WA during stance (*r*≈2). In contrast, in the CP group (figure 3), pelvic tilt exhibited SEM-BA greater than SEM-WA throughout the gait cycle (*r*≈1.5).

**Figure 1.**
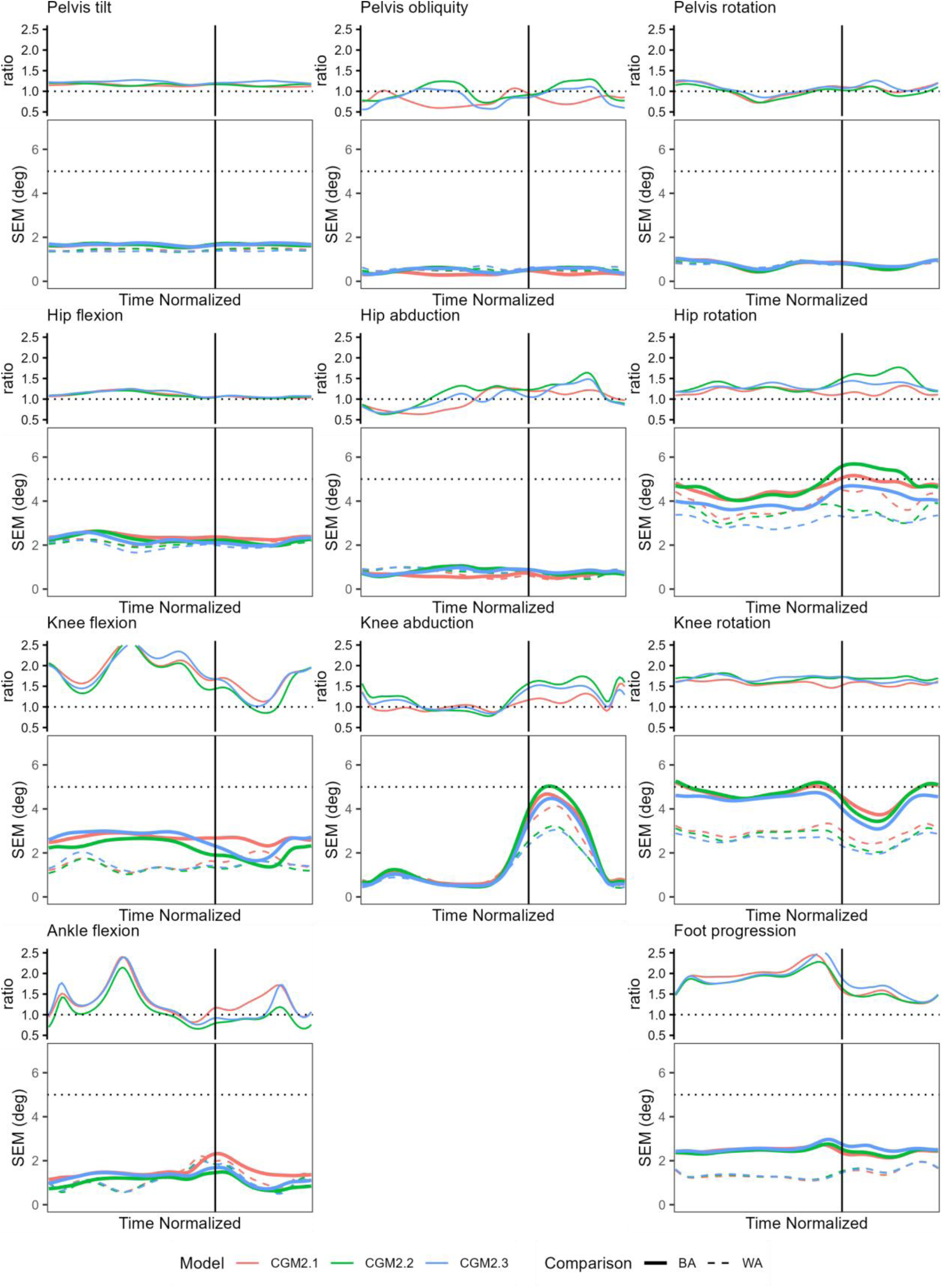
Time-normalized between-assessors (BA) and within-assessor (WA) standard error of measurement (SEM) and SEM-BA/SEM-WA ratio for the asymptomatic participants. SEM and ratio were computed for conventional gait models: CGM2.1, CGM2.2 and CGM2.3. The vertical black line represents foot-off on both ratio and SEM plots. The horizontal dotted lines represent the 5° threshold in SEM plots and the equality (ratio=1) in and ratio plots

**Figure 2.**
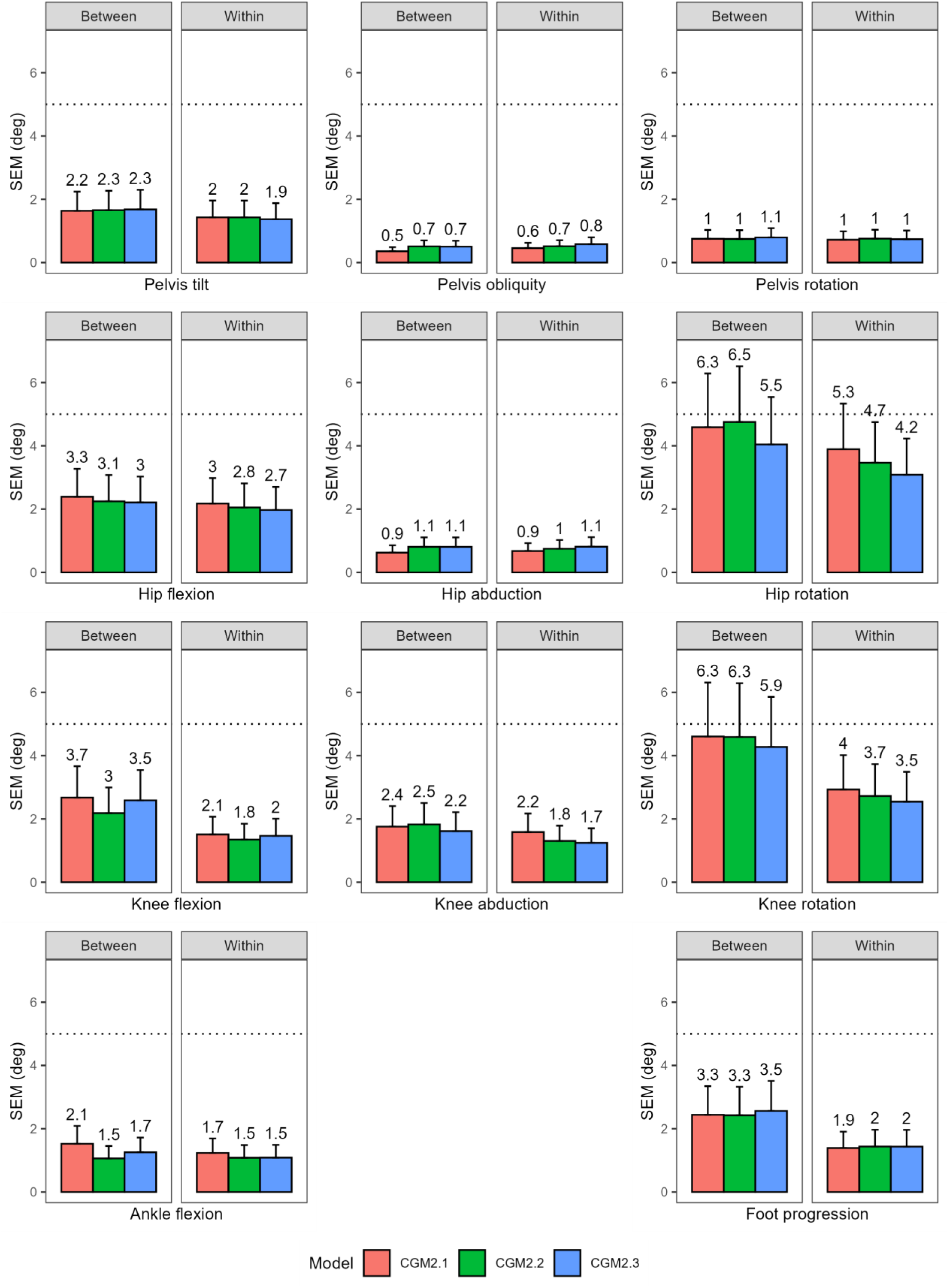
Average between-assessors and within-assessor standard error of measurement (SEM) of the asymtomatic participants for each joint angle of the conventional gait models: CGM2.1, CGM2.2 and CGM2.3. Value indicates the upper 95% confidence limit, calculated under the assumption that the variances follow a chi-squared distribution

**Figure 3.**
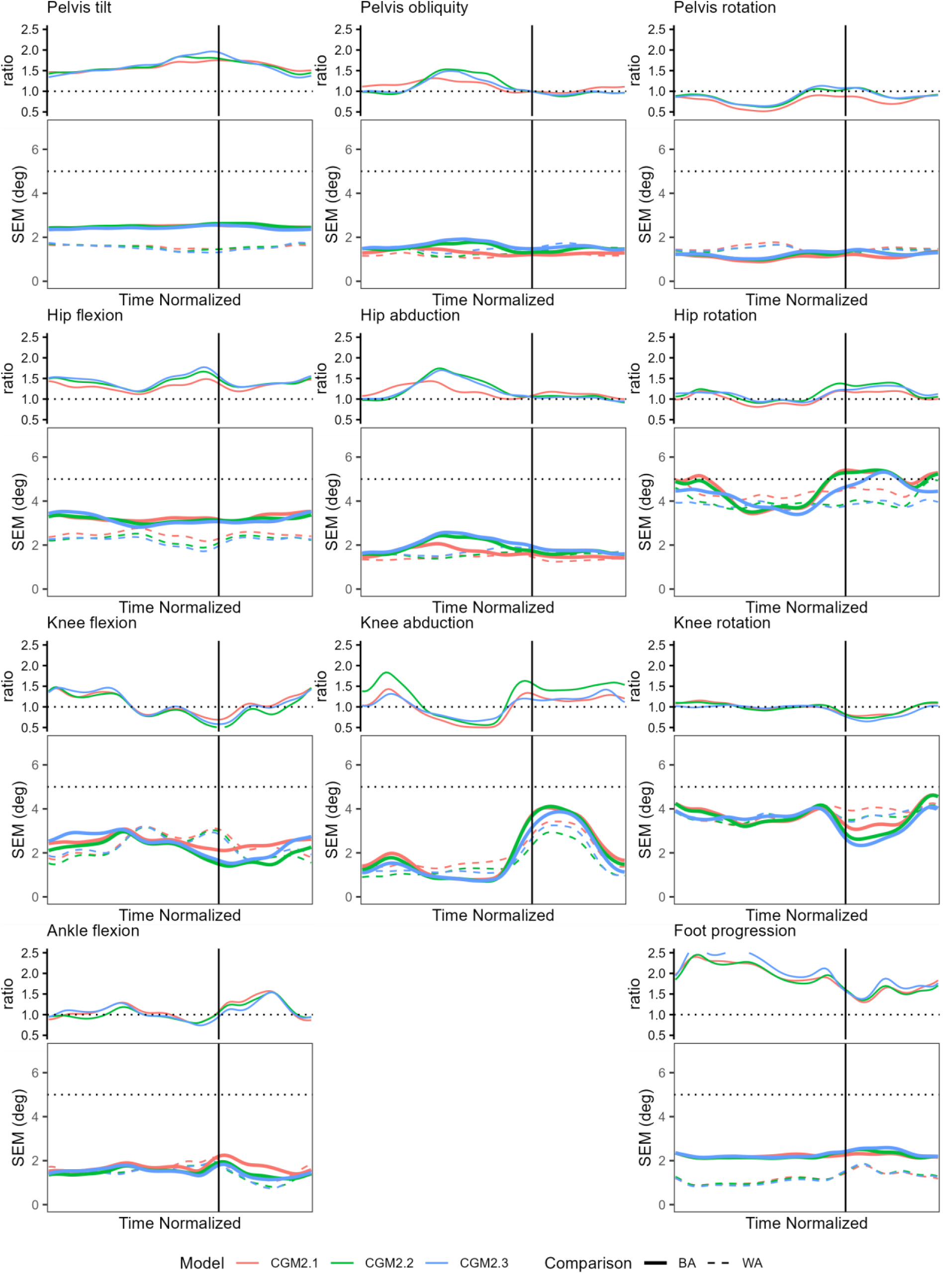
Time-normalized between-assessors (BA) and within-assessor (WA) standard error of measurement (SEM) and SEM-BA/SEM-WA ratio for the cerebral palsy participants. SEM and ratio were computed for conventional gait models: CGM2.1, CGM2.2 and CGM2.3. The vertical black line represents foot-off on both ratio and SEM plots. The horizontal dotted lines represent the 5° threshold in SEM plots and the equality (ratio=1) in and ratio plots

**Figure 4.**
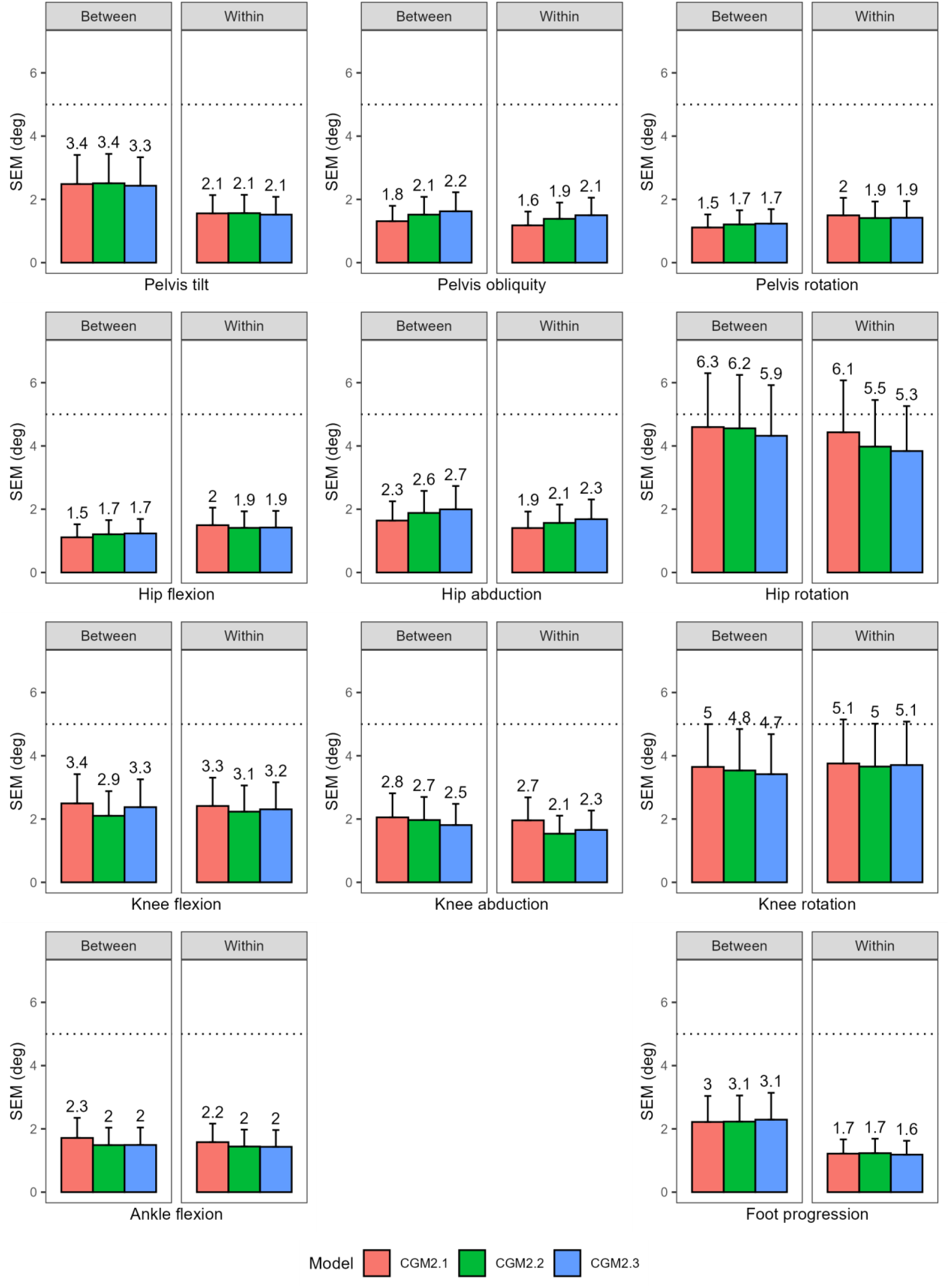
Average between-assessors and within-assessor standard error of measurement (SEM) of the cerebral palsy participants for each joint angle of the conventional gait models: CGM2.1, CGM2.2 and CGM2.3. Value indicates the upper 95 % confidence limit, calculated under the assumption that the variances follow a chi-squared distribution.

### 3.2. CGM2.1 and CGM2.2 comparison: the effect of the pose estimation method

The comparison between CGM2.1 and CGM2.2, using the same marker set but different pose estimation methods, showed that SEM exceeded 5° only for transverse plane rotations of the hip and knee. Around toe-off, SEM-BA for CGM2.2 was higher than for CGM2.1 in asymptomatic participants. The largest SEM-WA difference was 0.6° for hip transverse rotation in both groups. The largest difference SEM-BA was observed in knee flexion, with differences of 0.7° for asymptomatic and 0.5° for CP participants.

### 3.3. CGM2.2 and CGM2.3 comparison: the effect of additional markers on the thigh and shank

The comparison between CGM2.2 and CGM2.3, which differ by additional tracking markers on both thigh and shank, revealed similar effects across both groups, particularly for hip transverse rotation and knee flexion. For hip transverse rotation, the maximum SEM-BA decreased by 1° and the SEM-WA by 0.5° in asymptomatic participants with CGM2.3. This reduction persisted throughout the gait cycle. In the CP group, smaller reductions were observed, with decreases of 0.3° for between-assessor and 0.2° for SEM-WA.

CGM2.3 increased both SEM-BA and SEM-WA compared to CGM2.2 (by 0.5° and 0.4°, respectively) in both asymptomatic and CP participants. SEM for knee flexion with CGM2.3 were comparable to those observed with CGM2.1.

## 4. Discussion

The present study aimed to quantify the impact of methodological updates implemented across CGM2 iterations on the repeatability of kinematic outputs. Specifically, the transition from CGM2.1 to CGM2.2 assessed the influence of the segmental pose estimation method, whereas the transition from CGM2.2 to CGM2.3 evaluated the effect of the marker set modifications. Overall, both SEM-WA and SEM-BA remained around 2°, with MDC values near 6°, for both asymptomatic and CP groups. The use of kinematic fitting, as the default pose method in CGM2.2 produced only minor changes in SEM (maximum difference of 0.7° at the knee). In contrast, additional tracking markers in CGM2.3 yielded a modest reduction in hip transverse rotation SEM, approximately 1° across groups, suggesting a limited but positive effect of the updated marker set on repeatability. Together, these findings confirm that methodological refinements in CGM2 preserve the reliability of the original CGM.

Overall, both SEM-WA and SEM-BA were around 2°, and the MDC reached 6° for both groups and the various iterations of CGM2. These results align with those reported by Kainz et al.[26], Everaert et al.[27], and Wilken et al. [28], who respectively observed an SEM-BA of 3°, an SEM-WA of 2.9°, and an MDC of 5.8° in populations of children with CP and typically-developing children. In Kainz’s study [26], kinematic fitting was combined with rigid clusters to compensate for soft tissue artefacts. The comparable SEM observed in the present study, despite the use of skin-mounted markers, suggests that rigid clusters may not substantially improve repeatability, if markers are placed on an area less affected by the soft tissue artifact, i.e. the mid part of the segment [29]

In line with previous repeatability studies [26,27,30,31] and studies comparing biomechanical models [11,32] of gait, marker placement error primarily affects the transverse plane. Across CGM2 iterations and participant groups, SEM and MDC reached approximately 3° and 9° for the transverse plane. At the joint level, in the CP group, the SEM-BA and MDC-BA ranged from 4.3° to 4.6° and from 12° to 12.7°, respectively, for the hip rotation. Our CP group was primarily composed of participants with a GMFCS level 1 (n=15). However, lower MDC values than the ones reported by Klejman et al. [33] (between 18° and 23.4°) were observed in CP participants classified as GMFCS 1 and 3. In this current study, the focus was on analyzing the profile of SEM along the gait cycle rather than isolating a discrete value as done by Klejman et al. [33]. Aside from this methodological difference, the difference might also result from the intrinsic measurement errors of our CP participants, who might have been able to repeat gait trials consistently.

The maximum difference due to the pose estimation method, between CGM2.1 and CGM2.2, was 0.7° SEM for knee flexion. The constraints of kinematic fitting tend to reduce the error on hip transverse rotation, bringing SEM-WA below the 5° threshold [7].

Using CGM2.3, the error on hip transverse rotation was further reduced by approximately 1° with SEM-WA decreasing from 5.3° to 4.2° for asymptomatic participants and from 6.1° to 5.3° for CP, from CGM2.1 to 2.3, respectively. Hip transverse rotation is a major parameter in surgical decision-making, and reduced errors would better support clinical decisions. This reduction was also observed in Kainz et al. [26], where transitioning from the CGM to a musculoskeletal model using kinematic fitting reduced error in hip transverse rotation by 2° in their CP cohort. In general, reducing the number of joint degrees of freedom leads to lower errors as reported by Kainz et al. [26], Mentiplay et al.[34], and Duprey et al.[35]. One should exercise caution: constraining joint motion may improve repeatability, but may also lead to outputs that reflect model assumptions rather than true physiological motion. For instance, limiting the knee to a single degree of freedom would artificially reduce error in abduction/adduction or transverse rotation but would no longer capture deviations that may be clinically meaningful in pathological gait. CGM2.3 maintains three degrees of freedom at each joint, thereby preserving compatibility with the original CGM.

Anatomical marker misplacement remains the main source of extrinsic measurement errors [7,36]. Neither change in the pose estimation method nor modification to the marker set substantially affected kinematic outputs, with no apparent differences in errors between CGM2.2 and CGM2.3. Variability, therefore, mainly depend on the assessor’s training in anatomical palpation, as well as their ability to reproduce marker placement consistently; consequently, test–retest protocols remain essential for evaluating inter-assessor agreement. In our dataset, pelvis marker placement was less consistent in the CP group, whereas knee marker palpation showed greater errors in asymptomatic participants.

The main limitation of this study lies in the simplistic design of the test-retest protocol, involving only one session for one assessor and two sessions for the other assessor. Repeatability protocols are time-consuming and impose a substantial burden on patients, which limits their feasibility in clinical centers and makes data collection challenging. We opted for a protocol that could be adopted by most centres, reflecting the reality of clinical staff typically composed of a single assessor [37]. Additionally, no results on joint kinetics were reported. Finally, this study focused on measurement reliability rather than model accuracy; it does not provide evidence on which CGM2 iteration most accurately represents true bone motion.

This study showed that methodological updates across CGM2 iterations had minimal impact on the repeatability of kinematic outputs. The introduction of kinematic fitting in CGM2.2 preserved reliability, while additional tracking markers in CGM2.3 produced a small but consistent reduction in hip transverse rotation error. Overall, both SEM-BA and SEM-WA remained around 2°, confirming that CGM2 maintains the reliability of the original CGM. These results support the use of CGM2 in clinical gait analysis and highlight its compatibility with established CGM-based practice.

## Funding declaration

This study was funded by the Swiss National Fond (FNS), CRSII5_177179, (http://p3.snf.ch/project-177179).

Cloe Dussault Picard was funded by the Fonds de recherche du Québec —Nature et technologie for the postdoctoral scholarship.

## Author contributions Declaration

- **Fabien.Leboeuf.**: Conceptualization, Methodology, Software, Data curation, Formal analysis, Writing – original draft.
- **Morgan .Sangeux.**: Writing – review & editing, Supervision.
- **Mickael.Fonseca.**: Investigation, Writing – review & editing.
- **Cloé Dussault.-Picard.**: Writing – review & editing.
- **Stephane .Armand.**: Investigation, Writing – review & editing.

## Data availability statement

The datasets analysed during the current study are not publicly available due to institutional policy but are available from the corresponding author on reasonable request

## Notes

### Competing Interest Statement

The authors have declared no competing interest.

### Author Declarations

The study was approved by the commission cantonale ethique de la Recherche de Geneve

